# Prospective, Randomized, Parallel-Group, Open-Label Study to Evaluate the Effectiveness and Safety of IMU-838, in Combination with oseltamivir, in Adults with Coronavirus-19- a feasibility trial

**DOI:** 10.1101/2024.10.31.24316416

**Authors:** Kavi Sharma, Lisa Berry, Evangelos Vryonis, Asad Ali, Beatriz Lara, Angela Noufaily, Nick Parsons, Christopher Bradley, Becky Haley, Shivam Joshi, Tabuso Maria, Ramesh P Arasaradnam

**Affiliations:** University Hospitals Coventry & Warwickshire, Coventry, CV22DX; Warwick Medical School, Coventry, CV4 7AL; Brighton and Sussex Medical School, Falmer, BN1 9PH

**Keywords:** COVID-19, IMU-838, Vidofludimus calcium, oseltamivir

## Abstract

**Background:** The global urgency for effective treatments against SARS-CoV-2 infections, causing COVID-19, remains paramount. One promising avenue is repurposing existing medications. IMU-838, a dihydroorotate dehydrogenase (DHODH) inhibitor, has exhibited potent antiviral effects against respiratory viruses. In rodent studies, its combined administration with oseltamivir has shown therapeutic potential against both influenza and SARS-CoV-2.

**Objective:** The primary aim of the IONIC pilot feasibility trial was to comprehensively explore the feasibility and safety challenges associated with administering the novel treatment regimen of IMU-838 combined with oseltamivir to COVID-19 patients. The secondary objective was to evaluate whether a 14-day treatment course with IMU-838 and oseltamivir improves time to clinical improvement compared to oseltamivir alone, defined as the duration from randomization to achieving a 2-point improvement on the WHO ordinal scale, discharge from the hospital, or occurrence of death, whichever comes first.

**Methods:** IONIC was a Phase IIb, randomised, open-label, single centre trial. Prospective participants were recruited from a single centre within the UK and were eligible if they had moderate to severe COVID-19 requiring hospitalisation, were aged 18 years or above. Patients were randomly assigned, in a 1:1 ratio, to either the IONIC intervention arm (IMU-838 + oseltamivir + standard care) or the control arm (oseltamivir + standard care). Sponsored by the University Hospital Coventry & Warwickshire NHS Trust and funded by LifeArc, the trial comprises two distinct phases: a pilot feasibility study and a main study. The aim of the feasibility study was to inform the design and execution of the main study. However, due to recruitment challenges, the main study was not conducted. The present paper reports the results of the feasibility study. The trial was prospectively registered with ISRCTN (ISRCTN53038326) and Clinicaltrials.gov (NCT04516915)

**Findings:** Between 22 Jun 2020 and 20^th^ May 2022, 38 participants were recruited into the trial. Recruitment challenges hindered the main study, but the feasibility trial provided encouraging findings. Treatment completion rates stood at 84%, with no serious adverse effects observed affirming the safety profile of the novel treatment. Although no statistically significant difference emerged in time to clinical improvement between the treatment and control groups, logistic regression analysis indicated a notable association between the treatment group and clinical improvement within a 14-day window. These results emphasize the crucial role of feasibility studies in guiding larger-scale trials and underscore the necessity for further investigation into the therapeutic potential of this approach to the treatment of COVID-19.

## Introduction

The onset of the global pandemic declared by the World Health Organization (WHO) on March 11, 2020, in response to the severe acute respiratory syndrome coronavirus (SARS-CoV-2), underscored the urgency in combatting coronavirus disease 2019 (COVID-19). While COVID-19 typically manifests with symptoms such as fever, cough, fatigue, and dyspnoea, early meta-analyses suggested varying fatality rates, prompting an intense search for effective treatments(1) . At the commencement of the IONIC pilot/feasibility study reported here (in June 2020), no effective treatments for COVID-19 had been found. At this time, the anticipated severity of the epidemic indicated that hospitals, particularly intensive care units, may encounter substantial pressure, with various models of pandemic spread suggesting that up to half of the adult population could fall ill within 8-12 weeks without intervention, with roughly 10% requiring hospitalization(1–3). Leading to nearly 2 million hospital admissions in the UK alone. Considering this scenario, it was prudent to explore treatments that may offer only a modest effect on survival or on hospital resources(2).

The IONIC pilot/feasibility study represented a pioneering effort to assess the feasibility and challenges of implementing a novel treatment regimen combining IMU-838 and oseltamivir for COVID-19 hospitalised patients. Alongside, the study aimed to investigate whether this treatment course could expedite clinical improvement compared to oseltamivir alone.

## Choice of Intervention

IMU-838, also known as Vidofludimus, is a selective inhibitor of Dihydroorotate dehydrogenase (DHODH). Safety analysis of IMU-838 revealed no deaths or serious adverse events, with common side effects including headaches and gastrointestinal symptoms. IMU-838 shows promise in treating COVID-19 by inhibiting pyrimidine synthesis, disrupting viral replication, and activating the immune response(4–6). Studies suggest a synergistic response between IMU-838 and oseltamivir in treating influenza, and clinical trials are underway to evaluate their combined effect on COVID-19(7). Oseltamivir was chosen for its affordability and availability in the UK and Europe. Despite the lack of direct evidence for oseltamivir’s effectiveness against SARS-CoV-2, its combination with IMU-838 offers a cost-effective treatment option for COVID-19(8).

## Methodology

IONIC was a Phase IIb trial conducted as a randomised, open-label, investigation aimed at evaluating the efficacy of administering IMU-838 alongside oseltamivir to hospitalized COVID-19 patients, compared to oseltamivir alone, over a span of 14 days. The trial incorporated an internal pilot feasibility study mirroring all facets of the main study, intended to provide additional insights for refining the design and implementation of the main trial. The trial adhered to Good Clinical Practice and the Declaration of Helsinki guidelines, with ethics approval obtained from Wales Research Ethics Committee – (Ref No: 20/WA/0146). In addition, required regulatory approvals were received from the Medicines and Healthcare products Regulatory Agency (MHRA). The IONIC trial protocol has been published elsewhere(9).

Patients were included in the trial if they met all inclusion criteria and no exclusion criteria. The key inclusion criteria were; male or non-pregnant female patients (>18 years), having confirmed or suspected moderate to severe COVID-19 requiring hospitalisation. Exclusion criteria were known allergic or hypersensitivity to the IMU-838, Oseltamivir, or any of the ingredients, pregnant or breastfeeding or with intention to become pregnant during the study, medical or concomitant disease history preventing participation.

The trial comprised distinct phases, including a screening phase, a 14-day treatment period, a subsequent 14-day follow-up period, and a long-term follow-up extending up to one year. Throughout these phases, the study compared the effectiveness of the IONIC Intervention to the administration of oseltamivir alone. All participants received standard care, consistent with WHO recommendations, encompassing measures such as supplemental oxygen, antibiotic therapy, and vasopressor support. Post Day 14, patients continued with appropriate standard care as determined by the clinical care team. Follow-up data collection took place every four days, with long-term follow-up assessments scheduled at 3-, 6-, and 12-months post-enrolment.

Patients who were able to provide informed written consent were directly approached for enrolment into the study. In cases where patients lacked the capacity to provide consent, a relative or close acquaintance was approached for consent. Given restrictions on visitors in hospital settings, verbal consent was obtained via telephone and duly documented on the consent form.

In instances where neither a relative or a friend could be identified, a professional representative not affiliated with the trial, such as a doctor, could be approached to act as the legal representative. Subsequently, consent was sought from the patient’s personal legal representative, or directly from the patient if they recovered promptly, at the earliest feasible opportunity.

## Randomisation

Randomisation to treatment was implemented, at a 1:1 ratio, utilizing variable block sizes. Stratified randomization was facilitated through an online system, and the trial remained open label, ensuring both patients and staff were aware of treatment allocation. However, statisticians analysing outcome data remained blinded to treatment allocation.

## Objectives

The primary study objectives centred around evaluating treatment completion rates, protocol adherence, feasibility challenges, and safety profiles. This evaluation was based on the number of eligible participants screened, recruitment rates, documentation of adverse reactions, and participant compliance with the treatment regimen. The secondary objectives aimed to assess the time to clinical improvement, with exploratory endpoints encompassing effectiveness, safety, and long-term effects. The effectiveness of the IONIC Intervention was gauged in comparison to oseltamivir and standard care, focusing on the time-to-clinical improvement measured by a 2-point advancement on the WHO ordinal scale during the 14-day treatment period, which ranged from 0 to 8 (representing the spectrum from lowest to highest illness severity, with 8 indicating death). Participants were subsequently followed up either by phone or face-to-face, depending on their discharge status, on day 28 post-randomization, and their clinical status was recorded using the hospitalization clinical status scale (Table 1).

**Table 1:** Hospitalised clinical status.

|  |
| --- |
| Category 0 - No clinical or virological evidence of infection |
| Category 1 - not hospitalised with resumption of normal activities |
| Category 2 - not hospitalised but unable to resume normal activities |
| Category 3 - hospitalised no oxygen therapy |
| Category 4 - hospitalised, requiring supplemental oxygen by mask or nasal prongs |
| Category 5 - hospitalised, requiring non-invasive ventilation or nasal high-flow oxygen therapy |
| Category 6 - hospitalised requiring incubation or invasive mechanical ventilation |
| Category 7 - Ventilation + additional organ support - pressors, RRT, ECMO |
| Category 8 – Death |

## Trial Treatments

### IMU-838 (Vidofludimus calcium)

IMU-838 was administered twice daily as oral tablets starting with a loading dose of 45mg on the first day (Day 1, Table 2).

**Table 2:** Planned dosing scheme for IMU-838 in the IONIC study.

|  | Day 1 |  | Day 2-13 |  | Day 14 |  |
| --- | --- | --- | --- | --- | --- | --- |
| Time | AM | PM | AM | PM | AM | PM |
| Number of Tablets |  | 2 | 1 | 1 | 1 | 1 |
| Dose of IMU-838 |  | 45 mg | 22.5 mg | 22.5 mg | 22.5 mg | 22.5 mg |
| | | $\alpha$ | $\beta$ | $\beta$ | $\beta$ | $\beta$ |

$\alpha$ Day1: loading dose of 45mg IMU-838 once daily given on the evening of Day 1
$\beta$ Day 2-14: dosing 22.5 mg of IMU-838 BID

Day 2-14: Once in the morning (15-60 min before a meal), and once in the evening (at least 2 hours after any meal and 15-60 min before any meal).

## Oseltamivir

Oseltamivir was taken from commercially available stock with a UK Marketing Authorisation. Twenty-eight doses of oseltamivir 75mg were administered over 15 days as defined in Table 3 below.

**Table 3:** Planned dosing scheme for oseltamivir in the IONIC study.

|  | Day 1 |  | Day 2-14 |  | Day 15 |  |
| --- | --- | --- | --- | --- | --- | --- |
| Time | AM | PM | AM | PM | AM | PM |
| Number of Tablets |  | 1 | 1 | 1 | 1 |  |
| Dose of oseltamivir |  | 75 mg | 75 mg | 75 mg | 75 mg |  |
| | | $\alpha$ | $\beta$ | | $\gamma$ | |

$\alpha$ Day1: Single dose 75mg on the evening of Day 1 $\beta$ Day 2-14: dosing 75 mg of BID of oseltamivir $\gamma$ Day 15: Single dose 75mg on the morning of Day 15

## Statistical Analysis

Descriptive statistics were calculated for participant demographics, stratified by treatment arm. To evaluate the primary outcome of feasibility, we assessed several factors, including eligibility, recruitment, and consent rates, as well as rates of treatment adherence, tolerability, adverse events, follow-up, withdrawal, and discharge at 14 and 28 days for each arm. We also examined the feasibility of using the study’s outcome measures in a future larger-scale trial.

For secondary analyses, we compared the time to clinical improvement between the treatment arms using a proportional hazards survival model, adjusting for age, baseline clinical status, and comorbidities. Participants who did not achieve clinical improvement or who died within the 14-day period were right-censored. Hazard ratios with 95% confidence intervals were reported, and Kaplan-Meier survival curves were constructed to visualise the time to clinical improvement. Additionally, we fitted a logistic regression model to the binary outcome of clinical improvement, adjusting for age, sex, and comorbidities, with odds ratios and p-values presented. A sensitivity analysis was conducted to assess the impact of censored observations due to death on the overall conclusions. All analyses were conducted on an intention-to-treat basis using complete case analysis and performed using R (version 4.4.0).

## Results

Between June 2020 and May 2022, 1,111 potential participants were screened at a single centre (University Hospital Coventry & Warwickshire; UHCW) in the UK. Of these, 902 (81%) were deemed ineligible, primarily due to blood test results that were outside the acceptable range. Among the 209 (19%) potentially eligible participants, 92 (8%) were approached for consent, and 38 (3%) ultimately provided consent and were randomised into the study (Figure 1).

**Figure 1.**
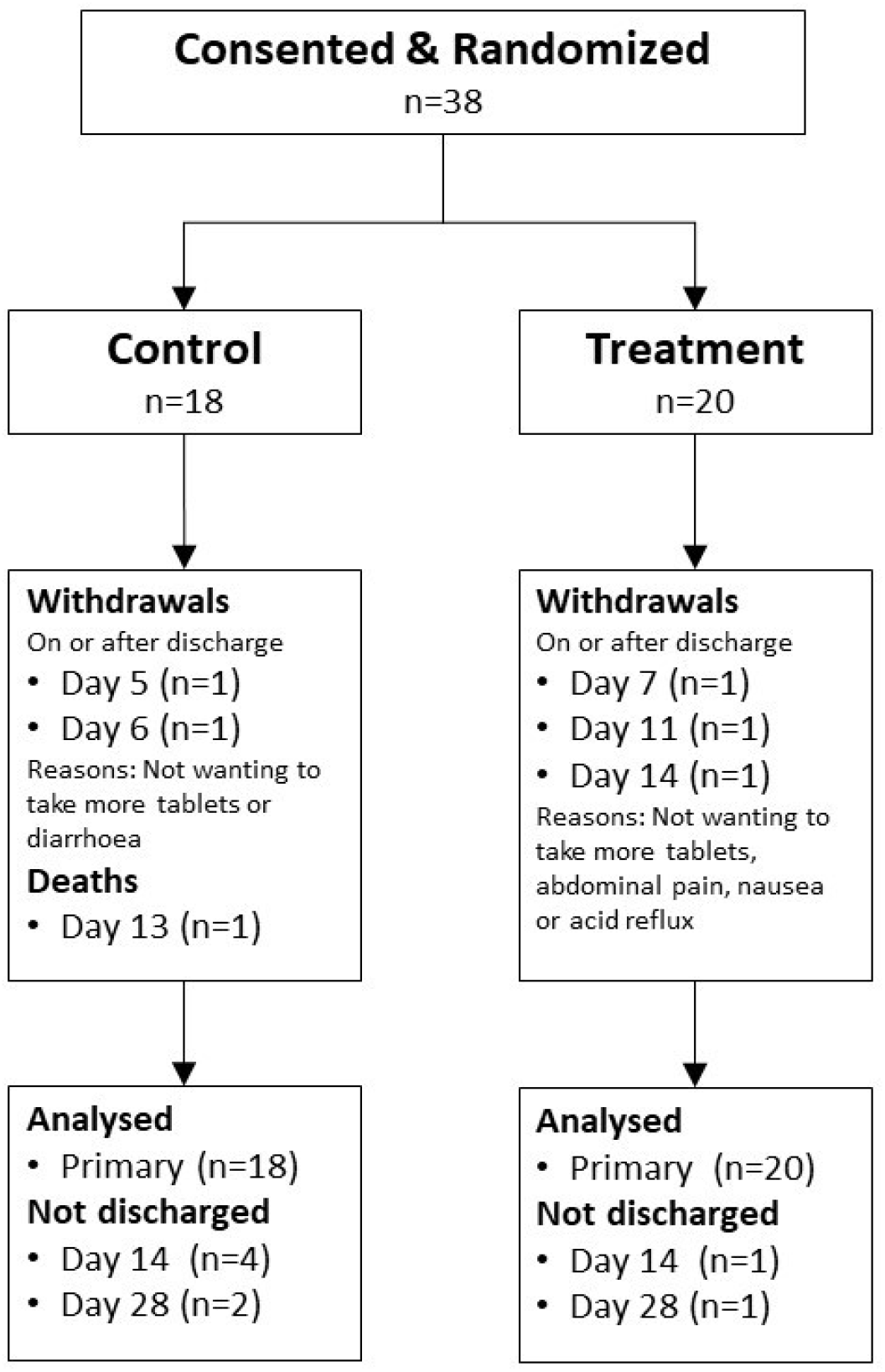
Participant Flow in IONIC trial

Several factors contributed to the low recruitment rate of eligible participants, including the prioritisation of participants for other trials, arrivals of potential participants outside regular working hours, and limited availability of medical staff (diverted to emergency care). Additionally, some potentially eligible participants were missed during the recruitment process. The trial began with a pilot/feasibility phase at UHCW to gather insights for the main study. However, despite achieving the recruitment targets for the pilot phase, it became clear that reaching the required sample size for the main trial would not be feasible through a single-centre approach.

## Participant Demographics

The baseline characteristics of the participants are summarised in Table 4. Of the 38 participants included in the analysis (mean age 56.4 years [SD 14.7]), 27 (71%) were male, and 24 (63%) were white British. Twenty participants were allocated to the treatment arm, while eighteen were assigned to the control arm. The mean age in the control group was 54.2 years (SD 14.4), with 13 (72%) males, whereas in the treatment group, the mean age was 58.4 years (SD 14.9), with 14 (70%) males.

**Table 4:** Baseline Data (at Hospital Entry) for IONIC Study Participants.

|  | **All participants**<br>**(n=38)** | **Control**<br>**(n=18)** | **Treatment**<br>**(n=20)** |
| --- | --- | --- | --- |
| **Age in years; mean (SD)** | 56.4 (14.7) | 54.2 (14.4) | 58.4 (14.9) |
| **Gender; n (%)** |  |  |  |
| **Female** | 11 (29%) | 5 (28%) | 6 (30%) |
| **Male** | 27 (71%) | 13 (72%) | 14 (70%) |
| **Ethnicity; n (%)** |  |  |  |
| **White British** | 24 (63%) | 8 (44%) | 16 (80%) |
| **Other** | 14 (36%) | 10 (56%) | 4 (20%) |
| **BMI; mean (SD)** | 30.4 (7.8) | 30.5 (9.4) | 30.4 (6.3) |
| **Oxygen saturation; mean (SD)** | 95.7 (2.0) | 95.3 (1.8) | 96 (2.2) |
| **Albumin; mean (SD) g/l** | 38.8 (4.0) | 38.9 (4.7) | 38.7 (3.2) |
| **CRP; mean (SD) mg/l** | 71.9 (56.4) | 61.9 (56.4) | 83.1 (56) |
| **Clinical Status; mean (SD)** | 3.9 (0.5) | 3.9 (0.5) | 3.9 (0.5) |
| **Patients with comorbidities** | 28 (73%) | 13 | 15 |
| **Patients with heart, lung, or diabetes** | 15 (39%) | 9 (50%) | 6 (30%) |

## Feasibility

### Treatment Adherence and Tolerability

The trial demonstrated strong treatment adherence, with 84% of participants completing the 14-day course.

### Adverse Events

Of the 38 participants, 21 (51%) reported adverse events, with 11 (52%) in the treatment arm and 10 (48%) in the control arm (Table 5). Most of the reported symptoms were mild to moderate, including nausea, constipation, palpitations, or COVID-19-related symptoms that were generally unrelated to the study treatment. In the treatment arm, four participants (19%) exhibited elevated Alanine Transaminase (ALT) levels, which were potentially related to the treatment, and one participant developed hospital-acquired pneumonia requiring ventilation due to COVID-19 complications. In the control arm, adverse events mainly included constipation, pulmonary embolism, pneumonitis, COVID-19-related complications, and numbness or weakness of the right arm or foot.

**Table 5:** Observed Adverse Events.

| **Adverse events** | **All participants**<br>**(n=38)** | **Control**<br>**(n=18)** | **Treatment**<br>**(n=20)** |
| --- | --- | --- | --- |
| *Number of participants with AEs* | 21 (51%) | 10 (48%) | 11 (52%) |
| *Total number of AEs* | 46 | 17 | 29 |
| *Nausea, Constipation, Palpitations* | 13 | 6 | 7 |
| *COVID related complications* | 3 | 2 | 1 |
| *High Alanine Transaminase (ALT)* | 4 | 0 | 4 |
| *Other* | 26 | 9 | 17 |

#### Withdrawal and Discharge

Five participants (13%) were not discharged within the 14-day follow-up period, including one from the treatment arm and four from the control arm. By day 28, three participants (8%) remained undischarged, with one from the treatment arm and two from the control arm (Table 6).

**Table 6:** IONIC Study Participant Follow-Up Data.

|  | **All participants**<br>**(n=38)** | **Control**<br>**(n=18)** | **Treatment**<br>**(n=20)** |
| --- | --- | --- | --- |
| **Withdrawals; n** | 5 (13%) | 2 (11%) | 3 (15%) |
| **Death; n** | 1 (2.6%) | 1 (5.5%) | - |
| **Not discharged within 14 days; n (%)** | 5 (13%) | 4 (22%) | 1 (5%) |
| **Not discharged within 28 days; n (%)** | 3 (8%) | 2 (11%) | 1 (5%) |
| **Outcomes** |  |  |  |

The findings suggest that the strong adherence to the treatment protocol, manageable adverse events, and low withdrawal rates indicate the feasibility of using “time to clinical improvement” as a primary outcome measure for future trials. The study demonstrated the potential for this outcome to be utilised effectively in a larger-scale clinical setting.

### Secondary Analyses

Secondary outcome data are summarised in Table 7. For participants who clinically improved or were discharged within the 14-day follow-up period, “time to clinical improvement” was defined as the number of days from the day of randomisation to the first follow-up day with a two-point improvement or discharge, whichever occurred first. For participants who were not discharged within 14 days, time to clinical improvement was set at 14 days. For those who died, it was calculated as the number of days from randomisation to the day of death.

**Table 7:** Time to Clinical Improvement for the IONIC Study.

|  | Control<br>(n=18) | Treatment<br>(n=20) | Difference in<br>means<br>(95% CI) | P-value* |
| --- | --- | --- | --- | --- |
| Time-to-clinical improvement for all participants (days); mean (SD) | 7.9 (4.1) | 6.6 (3.2) | 1.3 (-1.16, 3.74) | 0.292 |
| Time to clinical improvement for surviving participants discharged within 14 days (days); mean (SD) | 5.6 (1.9)<br>(n=13) | 6.2 (2.7)<br>(n=19) | -0.6 (-2.28, 1.09) | 0.477 |
| Time to clinical improvement for surviving participants (days); mean (SD) | 7.6 (4.0)<br>(n=17) | 6.6 (3.2)<br>(n=20) | 1.0 (-1.48, 3.46) | 0.420 |

\*P-values are based on independent samples t-tests.

**Table 8:** Logistic Regression.

| COVARIATE | OR (95% CI) | P-VALUE |
| --- | --- | --- |
| TREATMENT | 29.7 (1.22, 727) | 0.038 |
| **AGE** | 0.93 (0.85, 1.01) | 0.096 |
| **CLINICAL STATUS AT BASELINE** | 0.05 (0.00, 8.92) | 0.256 |
| **COMORBIDITIES** | 8.88 (0.41, 195) | 0.166 |

### Survival Analysis

The survival analysis compared the time to clinical improvement between treatment arms using a proportional hazards model adjusted for age, baseline clinical status, and comorbidities. None of these covariates, including treatment, age, clinical status at baseline, and comorbidities, were found to be statistically significant predictors of time to clinical improvement (hazard ratio for treatment: 0.70, 95% CI: 0.32–1.52, p = 0.366).

Figure 2 shows the Kaplan-Meier curves for both treatment arms, demonstrating that after 8 days, the proportion of participants achieving clinical improvement was lower in the treatment arm compared to the control arm. However, the log-rank test statistic comparing the survival curves was 3.58 (p = 0.500), indicating no statistically significant difference between the treatment groups in terms of time to clinical improvement.

**Figure 2.**
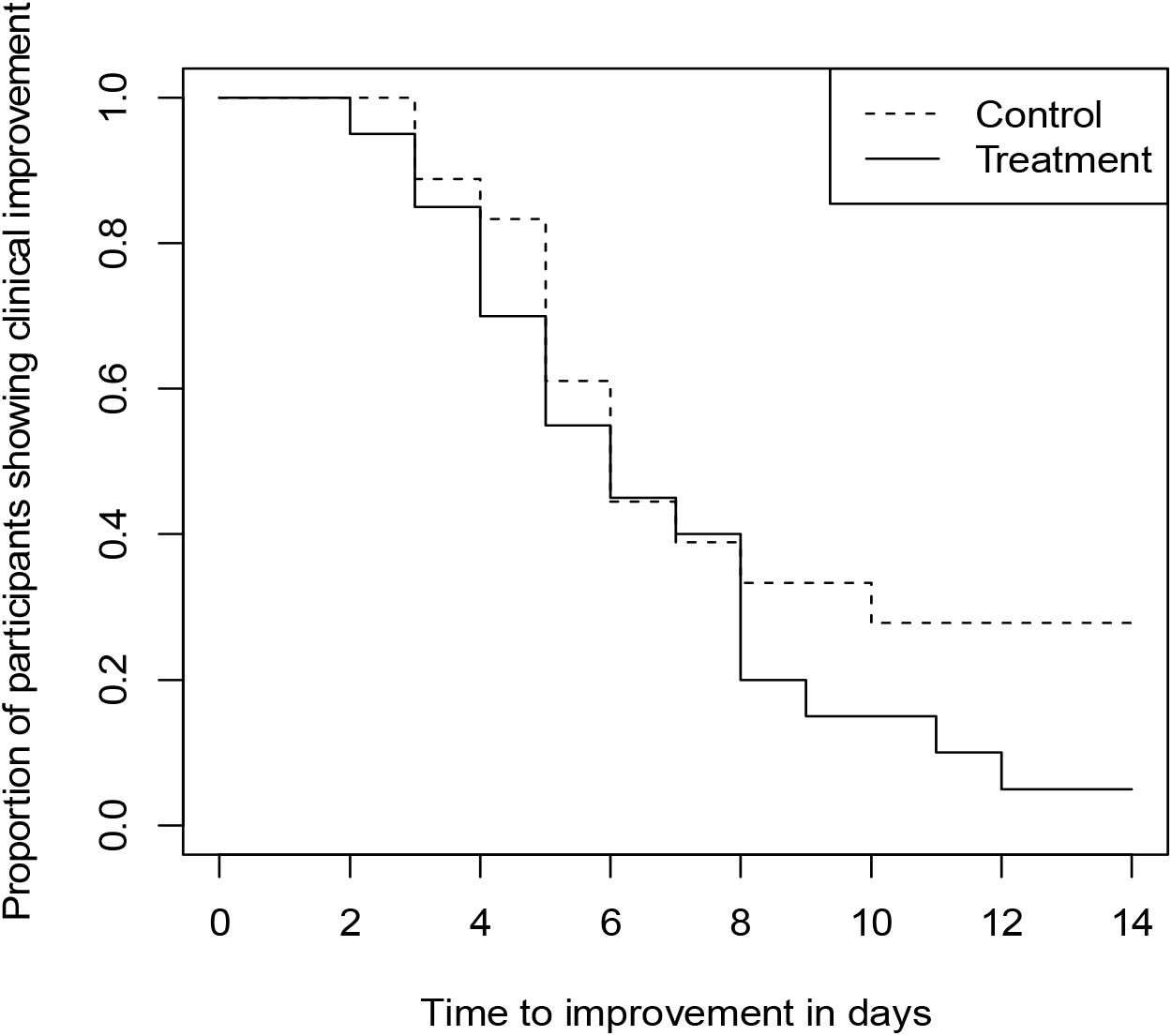
Kaplan-Meier Curves for Time to Clinical Improvement in the IONIC Study

### Logistic Regression

Logistic regression analysis examined the association between treatment and clinical improvement within 14 days, adjusting for age, sex, and comorbidities. The odds ratio for the treatment group was 29.7 (95% CI: 1.22–727, p = 0.038). However, the confidence interval is very wide, reflecting a high degree of uncertainty around this estimate, making it less reliable and difficult to interpret. Given this imprecision, the result should be interpreted with caution.

### Sensitivity Analysis

A sensitivity analysis was conducted to explore the effect of the censored observations due to death on the overall conclusions. This analysis assumed that the deceased participant had survived the treatment period but was not discharged by day 14. The results were consistent with the initial survival analysis, with a hazard ratio for the randomisation group of 0.698 (95% CI: 0.32–1.52, p = 0.366), suggesting that the initial findings were robust.

## Discussion

The IONIC feasibility trial aimed to evaluate the practicality of administering a novel treatment regimen combining IMU-838 with oseltamivir to hospitalised COVID-19 patients. A secondary objective was to assess the impact of this combination therapy on the time to clinical improvement. This trial contributes to the ongoing search for effective therapeutic strategies against SARS-CoV-2 infections, while also offering valuable insights into the feasibility and logistical challenges involved in delivering this combination therapy in a clinical setting.

### Feasibility of Recruitment and Study Implementation

Recruitment proved to be the primary challenge of the IONIC trial. Although we initially adopted a single-centre approach, recruitment difficulties necessitated a shift to a multicentre strategy. Factors such as the prioritisation of participants for other studies, logistical constraints (e.g., participants arriving outside of regular hours), and limited availability of medical staff significantly hindered enrolment. The COVID-19 pandemic further complicated recruitment efforts, disrupting healthcare systems and preventing the expansion to additional sites due to resource limitations, personnel shortages, and restricted hospital capacities.

While there was an attempt to extend recruitment to an international site in Ukraine, progress was halted due to the onset of the war, necessitating the suspension of these efforts. Ultimately, these challenges led to the decision to suspend the main trial to maintain the study’s integrity and avoid compromising research quality due to insufficient participant recruitment.

Moving forward, it will be essential to reassess the feasibility of conducting such trials in a post-pandemic context. Exploring alternative recruitment strategies, such as broader collaborations with other institutions or health networks, may help to overcome these obstacles and ensure future trial success.

### Participant Tolerability and Treatment Adherence

Despite these recruitment challenges, the feasibility trial demonstrated promising outcomes regarding participant adherence and tolerance to the treatment regimen. A strong adherence rate of 84% was observed, with most participants completing the 14-day course. Adverse events were documented, but the majority were mild to moderate and primarily related to COVID-19 complications rather than the treatment itself, thereby supporting the safety profile of the IMU-838 and oseltamivir combination therapy.

### Evaluation of Study Outcomes

One of the key secondary endpoints was the time to clinical improvement, defined as the duration from randomisation to the achievement of predefined clinical milestones. While the survival analysis did not reveal statistically significant differences in time to clinical improvement between the treatment and control groups, the logistic regression analysis suggested a potential signal of efficacy for the combination therapy within 14 days (odds ratio of 29.7, though this estimate was imprecise). However, the wide confidence interval indicates substantial uncertainty, which must be interpreted cautiously.

The study’s design, which used “time to clinical improvement” as a primary outcome, appears feasible based on participant adherence and the low rate of adverse events. However, the study’s limitations, particularly the small sample size, may have impacted the statistical power to detect a significant difference. The findings suggest that while there is a potential benefit of the combination therapy, further research with a larger sample size is needed to confirm these results and clarify the clinical relevance of this outcome measure.

### Conclusions and Recommendations for Future Research

In conclusion, the IONIC feasibility trial provides preliminary evidence supporting the feasibility of administering IMU-838 in combination with oseltamivir to COVID-19 patients, with a reasonable adherence rate and a manageable safety profile. Although the trial did not demonstrate a statistically significant difference in time to clinical improvement between the treatment arms, there was an indication of potential efficacy, which warrants further investigation.

Future studies would benefit from incorporating additional recruitment strategies and multicentre collaboration to enhance feasibility. These findings provide a foundation for designing larger-scale trials that could more definitively establish the therapeutic benefits of this promising combination therapy for COVID-19.

## Data Availability

All data produced in the present study are available upon reasonable request to the authors

## Author contributions

RPA and KS conceived of the presented idea to the funder. AA, LB and EV helped in developing the theory and delivery of the idea. NP and AN verified the analytical methods and the data analysis plan. LB and BL encouraged and assisted RPA to investigate specific of the trial including recruitment strategies, identification, and recruitment of the patients. KS has led on the project management with significant support from BH and CB. TM has led the research delivery team and assisted in recruitment. All authors discussed the results and contributed to the final manuscript.

## Acknowledgements

Mr John Todd^1^

Dr Neerja Bhala^2^, Prof Luca Frullon, Rosalba Radice,

Trial Management Unit (TMU), UHCW NHS Trust

National Institute of Health Research (NIHR) Coventry and Warwickshire Clinical Research Facility^3^

The clinical research delivery team

Research participants

This publication presents independent research funded by Life Arc & Immunic Therapeutics. The views expressed are those of the author(s) and not necessarily those of Life Arc, Immunic Therapeutics or the NHS.

## Footnotes

1 Patient and public representative

2 Independent chair IONIC Data monitoring Committee / Trial Steering Committee

3 This publication presents independent research funded by LifeArc and carried out with the support of the National Institute of Health Research (NIHR) Coventry and Warwickshire Clinical Research Facility. The views expressed are those of the author(s) and not necessarily those of LifeArc, the NHS, the NIHR or the Department of Health

